# Immunohistochemical Staining Characteristics of Well Differentiated Invasive Ductal Carcinoma Using the ADH5 Cocktail (CK5/14, P63, and CK7/18): a Potential Interpretative Pitfall

**DOI:** 10.1101/2022.11.16.22282316

**Authors:** Reham Al-Refai, Ahmed Bendari, Doaa Morrar, Sunder Sham, Sabina Hajiyeva

**Affiliations:** Lenox Hill Hospital, Northwell health, New York, NY, USA

## Abstract

**Context:** In our practice, an antibody cocktail ADH5 (CK5/14, p63, and CK7/18) helps with diagnostic challenges such as identifying microinvasion and foci of invasive carcinoma, differentiating atypical ductal hyperplasia from hyperplasia of the usual type and distinguishing basal phenotypes in triple-negative carcinomas. However, the ADH5 cocktail does have pitfalls and caveats.

**Objective:** To describe our experience with the ADH5 cocktail of antibodies in breast pathology.

**Data sources:** Institutional knowledge and literature search comprise our data sources.

**Conclusion:** The unexpected staining pattern of ADH5 in well-differentiated invasive ductal carcinomas can be challenging to interpret in these lesions with low-grade cytology. This occurrence, when using a multiplex stain, can confuse, and users should be aware of this potential pitfall.

## Introduction

Breast cancer is the most frequent non-skin cancer in women and the second leading cause of cancer death. Although all women are considered at risk, the risk varies by population. The lifetime risk of getting invasive breast cancer ranges from 3% (for women with no risk factors) to > 80% (for women with risk factors) (for women with highly penetrant germline mutations) (1).

According to the American Cancer Society, starting at the age of 40, all women have the option to start screening with a mammogram every year. Meanwhile, women 45 to 54 should get mammograms every year. Early identification is a significant component of increased survival after a breast cancer diagnosis. In specific patient populations, screening approaches for detecting breast lesions have improved, including the routine use of mammograms and ultrasound, as well as newer modalities such as magnetic resonance imaging (MRI) (2). The introduction of less invasive techniques resulted in smaller samples of lesional tissue becoming available for examination, and the ability to distinguish between benign, atypical, and malignant breast lesions, became firmly centered on microscopic features. Although detecting cancers early on may allow for more conservative surgical operations and increase the chance of a cure, diagnosing tiny tumors can be more difficult.

The mammary glandular system (ducts and lobules) comprises the inner layer of epithelial cells and the outer layer of myoepithelial cells. Such double-layered architecture is also present in epithelial proliferations (benign and atypical), sclerosing lesions, and in situ carcinomas. Consistent with their invasive nature, the glands in invasive carcinoma lack a myoepithelial layer. However, failure to detect myoepithelium is not independently diagnostic of invasive carcinoma. It can be lost in microglandular adenosis, some apocrine lesions, and some types of in situ ductal carcinoma.

Pathologists use an intact myoepithelial cell layer around cancerous cells as the most important diagnostic feature for differentiating in situ from invasive carcinomas (3). Diminished or absent myoepithelial stains in unequivocally benign lesions prompt the need to utilize at least two myoepithelial stains.

As a sensitive and specific marker, smooth muscle actin is one of the most commonly used stains. In addition to S100 protein, specific cytokeratins (keratins 5,7,14, and 17) also stain myoepithelial cells, but they are not very specific and not very sensitive (4). A p53 homolog, p63, has been identified as a selective marker of human epithelial basal cells in many organs, such as the skin, cervix, and prostate. Furthermore, p63 shows a high affinity for myoepithelial nuclei and is sensitive and specific to them in humans without concurrent staining for myofibroblasts, endothelial cells, pericytes, or smooth muscle cells. Unlike other markers, such as those that have been previously mentioned, p63 strongly stains myoepithelial cells’ nuclei, making them easily visible, whereas cytoplasmic markers are more difficult to discern. Staining fine-needle cytological preparations with p63 can also be beneficial when cytoplasm fragments occur during collection (5).

Grading of invasive ductal carcinomas provides an estimation of how closely it resembles normal breast glands. Well-differentiated carcinomas tend to be more slowly growing with a better prognosis. Tubular carcinoma is a special type of breast carcinoma with a favorable prognosis, composed of small, round to ovoid or angular glands and tubules with open lumens within a fibrous or fibroelastotic desmoplastic stroma. The cells that line the neoplastic tubules have relatively uniform nuclei of small or intermediate size. The diagnosis of tubular carcinoma requires that over 90% of the tumor consists of tubules and glands lined by a single layer of neoplastic cells. Tubular carcinomas are, by definition, well-differentiated carcinomas. The glands in tubular carcinoma have no surrounding myoepithelium, which helps differentiate them from benign conditions. The well-differentiated carcinoma cells have a luminal phenotype (CK8/18-positive, CK5/14-negative (6).

Breast core biopsy is one of the most common nonoperative methods of diagnosing breast lesions, and sometimes pathologists face difficulty giving as much diagnostic information as feasible in fewer tissue volumes (7,8).

ADH5 is a cocktail of antibodies targeting myoepithelial and epithelial antigens with dual chromogenic detection (brown chromogen: nuclear p63 and basal cytokeratins CK5 and CK14; red chromogen: luminal cytokeratins CK7 and CK18) is particularly useful when carcinoma present in biopsy material is limited. But the sensitivity of ADH5 for identifying well-differentiated carcinomas have not been investigated.

In the setting of foci suspicious for microinvasive carcinoma, to be able to colocalize the five antibody signals on one single slide are of utmost importance because the “suspicious areas” of microinvasive carcinoma is small and often might not be able to be detected on deeper cuts of tissue blocks.

Once invasive carcinoma of the breast is diagnosed, it is recommended that hormone receptor and HER2 testing be done on all primary breast carcinomas and recurrent or metastatic tumors (9); these are important prognostic and predictive markers.

The ADH5 cocktail is frequently utilized in our practice to help with diagnostic challenges such as identifying microinvasion and foci of invasive carcinoma, differentiating atypical ductal hyperplasia from hyperplasia of the usual type, and distinguishing basal phenotypes in triple-negative carcinomas.

Being widely used by pathologists within our group, an unexpected staining pattern was repeatedly observed in our practice when the ADH5 cocktail stain was used to prove the invasive nature of well-differentiated invasive breast carcinoma of no special type (ductal), and more so in tubular carcinomas, prompting further investigation into the issue. Most well-differentiated invasive breast carcinomas belong to the luminal A intrinsic molecular subtype and are positive for hormone receptors and negative for HER2.

Here, we aimed to review the ADH5 cocktail staining of well-differentiated invasive breast carcinoma of no special type (ductal), and tubular carcinomas, and compare it to a single stain of other myoepithelial markers (p63 and SMMH) to assess interpretative ease and ability to provide diagnostic information.

## Methods

### 1. Cases

After obtaining board approval from Feinstein Institutes for Medical Research/Northwell Health system, located in New York City, USA, we identified consecutive in-house breast lumpectomies and mastectomies diagnosed with well-differentiated invasive breast carcinoma of no special type (ductal) and tubular carcinomas between January 2017 and December 2021. A total of 44 consecutive cases were selected, and the diagnoses of all patients were confirmed by reviewing all hematoxylin and eosin (H&E) and IHC stained sections pertinent to the case by the authors, including our subspecialized breast pathologist (SH). Twenty-eight lumpectomies and 16 mastectomies were included in the study. Clinicopathologic information was obtained from pathology reports.

### 2. Immunohistochemistry

Unstained slides were prepared from the appropriate paraffin block of each case and stained with ADH5 cocktail (CK5/14, P63, and CK7/18) and smooth muscle myosin heavy chain (SMMH) using Ventana Benchmark Ultra System after antigen retrieval was applied.

For ADH5 cocktail: Predilute primary antibody (AVI3204) is applied to tissue sections for 32 minutes at 37 C, followed by Ultraview DAB detection kit. Then, denaturation for 4 minutes at 90 C. Followed by the Roche Ultraview universal alkaline phosphatase red detection Kit. Slides are then counterstained with hematoxylin, dehydrated (air dry and xylene), and coverslipped. Normal breast ducts and lobules were used as positive control.

For SMMH staining: After sections are backed and deparaffinized, A primary antibody SMMS-1 is applied for 16 minutes at 37 C. Visualization of antigens is achieved with an I-View DAB detection kit. Slides were then counterstained with hematoxylin, dehydrated (air dry and xylene), and coverslipped. A normal large intestine was used as a positive control. Table 1 shows the clone, dilution, pretreatment, and vendor for each antibody.

**Table1.** Clone, Dilution, Pretreatment, and Vendor of Antibodies Used for stains we used.

| Antibody | Clone | Pretreatment | Isotype | Type | Vendor |
| --- | --- | --- | --- | --- | --- |
| Anti-CK 5/14 | 34βE12 | Ultra CC1<br>standard (64 min)<br>at 95°C. | IgG1/<br>kappa | Mouse<br>monoclonal | Biocare<br>medical/ CA |
| Anti-P63 | 4A4 |  | IgG2a/<br>kappa | Mouse<br>monoclonal |  |
| Anti-CK7 | BC1 |  | IgG | Rabbit<br>monoclonal |  |
| Anti CK18 | 5D3 |  | IgG1 | Mouse<br>monoclonal |  |
| Anti-SMMH | SMMS-1 | CC1 (36 min) at<br>95°C | IgG1/<br>kappa | Mouse<br>monoclonal | Cell<br>Marque/Sigma<br>-Aldrich<br>company/CA |

### 3. Review of the morphology

Three authors studied H&E and the corresponding IHC slides for the ADH5 cocktail and single stains for P63, SMMH, and staining characteristics in carcinoma and surrounding normal ducts.

In both intraductal/luminal epithelium and myoepithelial cells, positive staining for CK5/14 was identified as brown cytoplasmic staining. In the cells of the intraductal/luminal epithelium, positive staining for CK7/18 was found as red cytoplasmic staining. Myoepithelial cells stained positive for p63 by showing brown nuclear staining and stained positive for cytoplasmic SMMH Table 2.

**Table 2:** The immunohistochemical stains that we used.

| Antibody | Staining | Cell type | Stain localization |
| --- | --- | --- | --- |
| Anti-CK 5/14 | DAB<br>Brown | Myoepithelial/Luminal Basal Phenotype | Cytoplasmic |
| Anti-P63 | DAB<br>Brown | Myoepithelial cells | Nuclear |
| Anti-CK 7/18 | FR<br>Red | Luminal epithelium | Cytoplasmic |
| Anti-SMMH | DAB<br>Brown | Myoepithelial cells | Cytoplasmic |

The immunostained slides were evaluated on the following criteria: visual ease of interpretation (based on the chromogen and antibody distribution) and amount of diagnostic information provided and were recorded as follows: expected and unexpected patterns.

## Results

### 1. Characteristics of the specimen and its morphology

The findings are summarized in Table 3. Of these 44 cases, 38 were tubular carcinomas and 6 well-differentiated invasive breast carcinoma of no special type (ductal). Of the 44 invasive carcinomas, 16 cases were identified in mastectomy specimens, whereas the remaining 28 were present in lumpectomy specimens. The histologic characteristics of in situ carcinoma were not further evaluated for this study.

**Table 3.** Characteristics of Breast Carcinoma Highlighted with the ADH5 Stain Abbreviations: ER: estrogen receptor, PR: progesterone receptor, HER2: human epidermal growth factor receptor 2.

| Clinicopathologic characteristics | No. | % |
| --- | --- | --- |
| Median age: |  |  |
| - 40-49 | 7 | 15.9% |
| - 50-59 | 12 | 27.2% |
| - 60-69 | 8 | 18.1% |
| - 70-79 | 14 | 31.8% |
| - 80-85 | 3 | 6.8% |
| Specimen type: |  |  |
| - Lumpectomy | 28 | 63.6% |
| - Total mastectomy | 16 | 36.3% |
| Specimen laterality: |  |  |
| - Right | 17 | 38.6% |
| - Left | 27 | 61.3% |
| Size of the lesion: |  |  |
| - 0.1-0.9cm | 22 | 50% |
| - 1.0-1.9cm | 17 | 38.6% |
| - 2.0-2.9cm | 3 | 6.8% |
| - 3.0-3.5cm | 2 | 4.5% |
| ER status |  |  |
| - Positive | 44 | 100% |
| - Negative | 0 | 0% |
| PR status |  |  |
| - Positive | 38 | 86.3% |
| - Negative | 6 | 13.6% |
| HER2 status |  |  |
| - Positive | 0 | 0% |
| - Negative | 44 | 100% |
| ADH5 staining |  |  |
| - Expected | 40 | 90.9% |
| - Unexpected | 4 | 9.1% |

### 2. Immunohistochemistry

We analyzed a total of 44 cases, 4 cases were previously (at the time of diagnosis) stained by the ADH5 cocktail and 40 cases we retrospectively stained to assess the usefulness and ease of interpretation of such multiplex stain in previously diagnosed invasive carcinoma. Four out of a total of 44 cases (9.1%), two tubular carcinomas and two well-differentiated invasive breast carcinoma of no special type (ductal), showed an expected pattern of staining for ADH5 with a loss of brown (P63, CK5/14) staining around invasive glands and diffuse red (CK7/18) expression Fig1-B.

**Fig.1.**
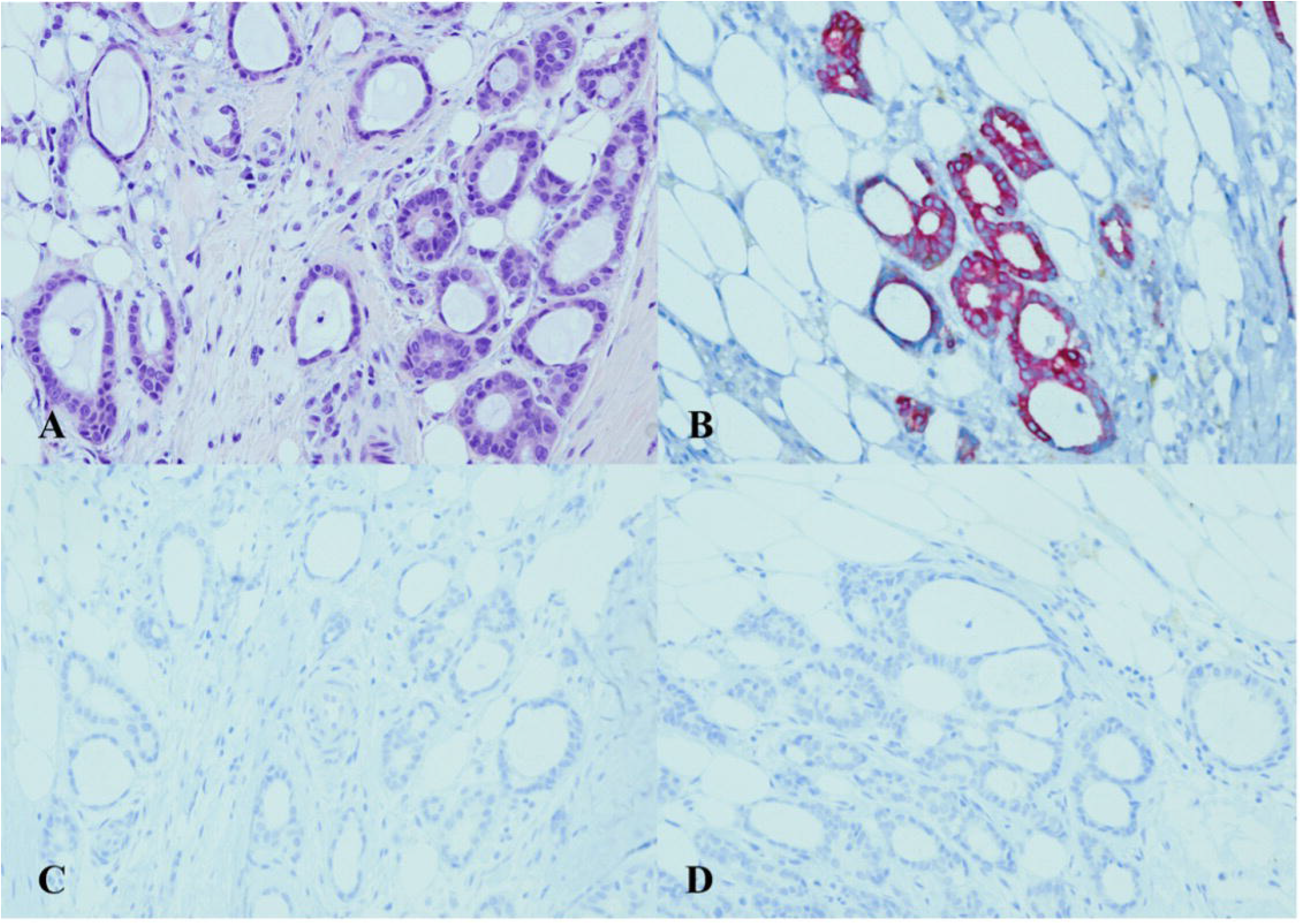
ADH5 Stain shows the expected pattern of staining. Invasive carcinoma shows a glandular growth pattern and a low nuclear grade with desmoplastic stroma (H&E) (**A**, ×20). The cytokeratin cocktail Stain confirms the presence of invasive ductal carcinoma (CK7/18 red) (**B**, ×20). The absence of corresponding myoepithelial stains (loss of brown P63 and CK5/14) confirms the invasive carcinoma (**C, D** ×20).

Forty out of 44 (91%) cases showed an unexpected staining pattern (mixture of cytoplasmic brown and red) (Fig.2-B).

**Fig.2:**
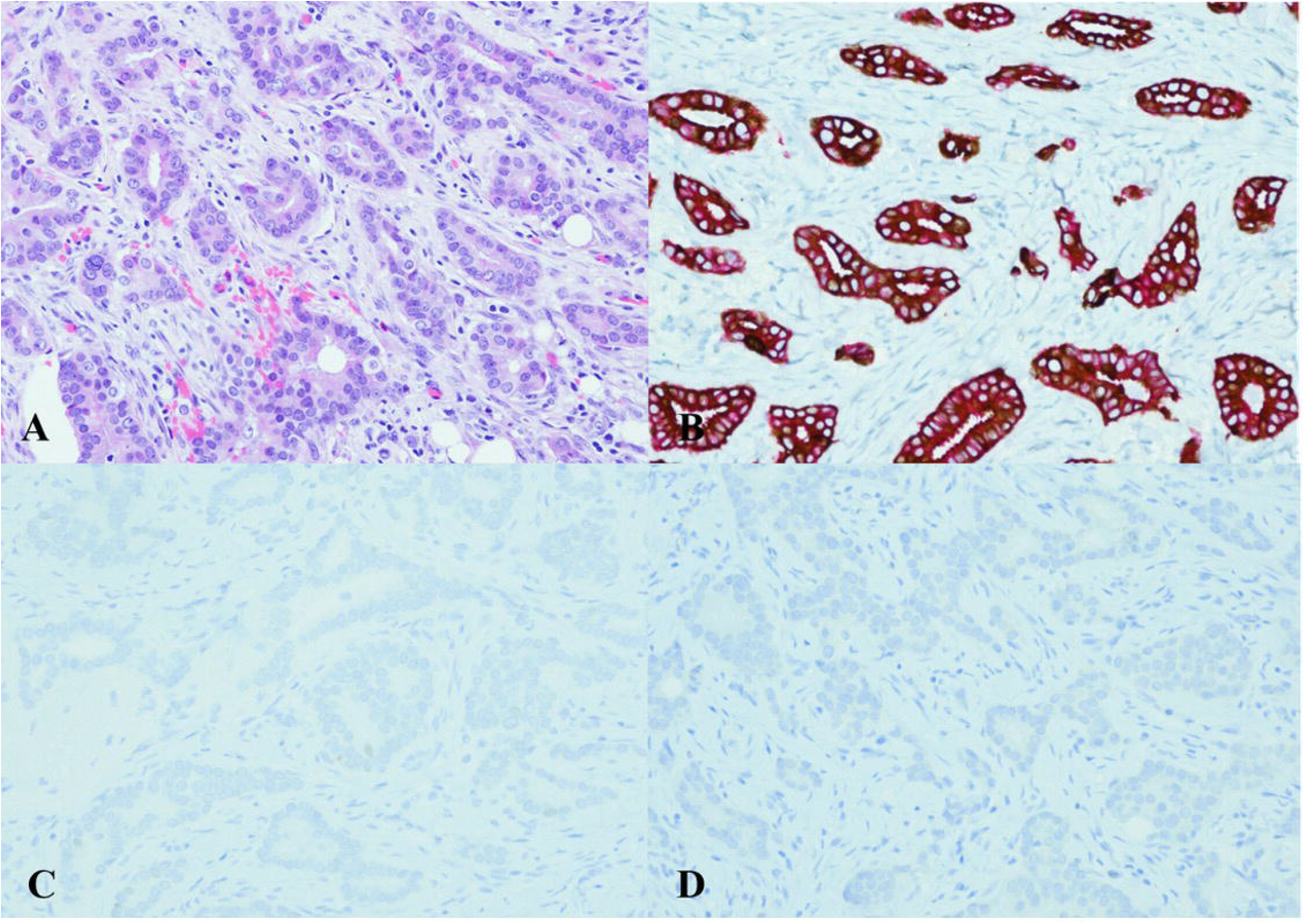
ADH5 Stain shows an unexpected pattern of staining. Invasive carcinoma shows a glandular growth pattern and a low nuclear grade with desmoplastic stroma (H&E) (**A** ×20). The cytokeratin cocktail shows the invasive ductal carcinoma foci staining mixed cytoplasmic brown and red (**B** ×20). The corresponding additional single myoepithelial stains confirm the loss of myoepithelial stains (loss of P63 and SMMH) (**C, D** ×20).

All 44 cases (100 %) showed negative myoepithelial staining for P63 and SMMH around invasive foci. (Fig .1-C, D and 2-C, D)

## Discussion

In the breast, there is a spectrum of ductal lesions from benign (usual ductal hyperplasia), borderline (atypical ductal hyperplasia), and preinvasive (ductal carcinoma in situ) to invasive (invasive ductal carcinoma). As these conditions are managed differently, the distinction between them is of critical clinical importance (10,11).

According to molecular and immunohistochemical studies, breast cancer is classified as basal-like, luminal A and B subtypes. “Basal-like” term used when tumors have high expression of genes showing features of basal epithelial cells of the normal mammary glands, including CK5/6, CK14, CK15, and CK17, and these cancers are usually triple-negative (ER-, PR-, HER2). Luminal subtypes include luminal A (ER+ [strong], PR+, HER2-) and luminal B (ER+ [weak / moderate], PR-, sometimes HER2+). Luminal cytokeratins are CK7/8, 18 and 19 (12,13,14,15,6).

Basal-like cancers are associated with high-grade morphology. Luminal A breast cancers are usually low grade, with slow growth and a better prognosis than luminal B cancers (6). Invasive carcinoma can generally be detected based on morphology alone; however, in challenging cases, using a variety of immunohistochemical markers that show the presence or absence of myoepithelial cells can be extremely helpful. Although P63 is frequently the stain of choice for myoepithelial cells, in situ carcinomas, and normal breast tissue, p63’s nuclear staining pattern results in an incomplete outer rim. As a result, attenuated myoepithelial cells can be more clearly observed when the nuclear p63 stain is combined with a cytoplasmic stain, such as that found with high molecular weight CKs (16,3).

In prior years, the number of immunohistochemical stains at our disposal was limited, but today we have several markers for the detection of myoepithelial cells, such as alpha-SMA, SMMH, Caldesmon, S100, P63, and others.

The increasingly more common limitation of the biopsy material is the amount of lesional tissue available for additional studies. In cases of breast carcinoma, there are further considerations even after the diagnosis because assessing biomarkers (estrogen/ progesterone receptors and HER-2/neu) is necessary to guide further clinical management and treatment (17).

The ADH5 cocktail stain (p63/CK7/18/CK5/14) is a valuable tool in breast pathology diagnostics, assessing potential invasion, and distinguishing between typical hyperplastic and atypical ductal epithelial proliferation. Furthermore, when employed in the minute foci of invasion setting, it appears to have the most significant diagnostic usefulness compared to single myoepithelial markers like SMMH and P63 (18). A similar concept of multiplex immunohistochemistry is applied in other organ systems such as the lung, brain, and prostate (for example, PIN4) (19,20,21).

Many issues have been encountered with multiplex stain, from technical to interpretative faults (22,23,19). In breast pathology, some studies looked into tissue integrity, quality of staining, and ease of interpretation for three multiplex stains (Minimal Carcinoma triple stain (MC), Breast Triple Stain (BTS) and LC/DC Breast Cocktail (LCDC). These stains are used to differentiate intraepithelial ductal proliferation and clarify the presence (and extent) of invasive carcinoma from in situ carcinoma or classify ductal or lobular proliferation (17).

We aimed to evaluate the diagnostic utility of the ADH5 cocktail stain by assessing the visual ease of interpretation (based on chromogen and antibody composition) in well-differentiated ductal carcinomas. We have repeatedly observed an unexpected staining pattern in well-differentiated ductal carcinomas in our practice and an investigation into such occurrences was necessary.

After reviewing a positive internal control, indicating all samples were appropriately processed and stained. We found 91% (40/44) of cases with unexpected staining patterns (a mixture of cytoplasmic brown and cytoplasmic red) instead of the expected pattern with loss of brown staining for (P63, CK5/14) around invasive glands and diffuse red staining for (CK7/18).

Such occurrence can happen in multiplex stains, though it is crucial to be aware of such potential pitfalls (7). Therefore, relying solely on immunohistochemistry can lead to a misdiagnosis. Many pitfalls of immunohistochemistry encountered in breast pathology strongly emphasize the importance of careful histopathological evaluation (24). An unexpected ADH5 staining pattern in invasive carcinomas, along with the low-grade cytological atypia may give the impression of benign glands. A multiplex stain can be misleading in cases of well-differentiated invasive ductal carcinoma, and users should be aware of this possibility.

## Data Availability

All data produced in the present study are available upon reasonable request to the authors

https://academicworks.medicine.hofstra.edu/academic_competition/2022/clinical_science/66/

## Ethical

ethical approval is waived..

## Patient Consent

Not required.

